# Early Warning Model for Patient Deterioration: A Machine Learning Approach for Nurse-Led Monitoring

**DOI:** 10.1101/2025.06.20.25329978

**Authors:** Esmael Ahmed, Mohammed Omer, Nuru Endris

## Abstract

The early recognition of clinical deterioration in hospital inpatients continues to be a major challenge in healthcare. In this work, we proposed an intelligible machine learning (iML) based EWS for predicting patient deterioration events and facilitating early nurse interventions. We compare a range of supervised learning models, including gradient boosting and logistic regression on electronic health record (EHR) data, emphasizing predictive performance and model interpretability. To be more clinically trusted and usable, we include an integration of SHAP (SHapley Additive exPlanations) for feature attributions and inline interpretability in alert interfaces. Our findings show that the proposed system not only has high predictive performance but also has a significantly positive impact on the nurse response behavior upon the actionable and interpretable alert generation. Transparency and user-centered design are further emphasized as keys to encouraging adoption in clinical practice. These results are a step toward the larger goal of incorporating AI into healthcare processes in a way that does not erode safety, trust, or human supervision.

**Author summary:** Monitoring patient decline is a crucial but serious problem in hospitals as it should be detected as early as possible. This work aimed to design an interpretable machine learning (iML) model to predict clinical deterioration based on patient electronic health records. In contrast to the classical black-box model, our system is highly accurate, transparent, and can be used by frontline healthcare professionals. We tested several machine learning methods and added SHAP (Shapley Additive exPlanations) to provide insight into how we make predictions. These explanations are on the alert interface, and this makes it easier to understand and trust the system by the nurses. Our findings demonstrate that the alerts enable nurses to react faster and better to care that resulting in better care coordination. This study emphasizes the need to develop AI tools that complement the clinician’s judgment, rather than substitute it, and that are simple to add to the existing hospital procedures. Our framework presents a solution in enhancing patient safety via AI without sacrificing the aspects of human control and confidence.

## 1 Introduction

In hospitals, it is still challenging for nurses to promptly notice if a patient is worsening. Some recent studies have found that up to half of known sepsis and respiratory complications are missed during routine checking [1]. Delayed diagnosis for a bloodstream infection increases the risk of serious health events, causing around 2 in 18 hospital deaths. Besides affecting patients, delayed recognition of deterioration results in patients staying at the hospital for an average of 3.2 extra days, causing the hospital extra expenses of $8,700 [2]. Since nurses are in constant contact with patients, they could notice early warning signs, but most systems do not help them enough in this task.

Despite being used extensively, traditional Early Warning Scores (EWS) like MEWS and NEWS are limited by three main problems that hinder their effectiveness. As key vital statistics in a hospital depend on paper tracking by nurses, nurses are often tied up for a long time during shifts checking, counting, and writing down vital signs [3]. Second, these systems report more false positive alarms than correct ones (over 70% in general wards), and this leads to a decline in how useful they seem. The static threshold approach does not consider a patient’s baseline or additional information from the nursing notes. The biggest challenge, it seems, is that they treat data separately rather than looking at everything hospitals have, including active monitor readings, notes from nurses, and lists of medications given to patients, which can all reveal helpful signs of deterioration [3].

Machine Learning(ML) approaches allow for a shift in deterioration prediction by examining detailed and regularly accumulating medical records at the same time. Modern neural networks can parse continuous data from wearables and nursing documents that contain unstructured text [4]. In the ICU, using models like LSTMs has proved to be good at detecting declines in health 4-6 hours before the symptoms appear. They work differently from fixed EWS, as they are aware of each patient’s norms and adjust to new medicine or symptoms mentioned by family members. Examining MIMIC-IV suggests that using both nursing notes and vital signs in the model boosts the AUC by 0.12, suggesting that nurses add important qualitative information not caught in the structuring fields [4].

Clinical data in deterioration prediction tends to have an overwhelming number of negative cases compared to positive deterioration observations. Our previous work on using advanced methods for imbalanced data and cost-sensitive learning in employee attrition prediction matches this challenge perfectly. Our proposal is to combine SMOTE with attention-based networks to ensure learning targets the most critical periods in time. To counter this issue, strong bias reduction measures are used along with this invention, since previous reviews uncovered unfair differences in predictions for people who speak another language or have a different form of a disease [5].

The critical evaluation of an AI system in healthcare happens when frontline nurses use it, and this requires handling three important human factors. Our work focuses on visualizing the SHAP values in terms that make sense to nurses [6]. Furthermore, to be useful, alerts need to fit into ongoing processes that guide our creation of stand-by dashboards and what is included in shift change briefings. In addition, the system ought to reflect a real-time benefit; our pilot results show nurses might reclaim 18 minutes per shift through automation. By including nursing staff in the development process, we address what truly matters and prevent any unnecessary use of technology. This differs from previous medical approaches since it recognizes that nurses are more likely to detect 73% of early warning signs [7].

In this research, an understandable machine learning model is built and confirmed to assist nurses in monitoring patients better by processing different types of clinical information to identify potential patient deterioration. Its study measures the impact the model has on nursing duties and judgments, using metrics that focus on nurses.

AI for clinical deterioration prediction has changed a lot over several important stages. Initial work in the area showed that machine learning can be used for risk identification, as logistic regression models based on vital signs were found to perform better than conventional risk scores in the ICU (AUC 0.82, better than 0.74). Research expanded to care given on general hospital wards and [8] was able to predict results across many hospitals with gradient boosted trees, while [9] included lab values for 4-6 hour predictions in advance. But, the methods used to begin with overlooked examples of nursing records that show when someone was getting worse [10].

Many important improvements were made through the use of temporal modeling techniques. With LSTM, it became possible to follow vital sign trends, which resulted in AUC increases of 0.88 to 0.92 in past studies. Improved results were discovered in transformer architectures [11] by using long-range information, but these architectures still faced generalization problems when being used in many hospitals (AUC decreased by 0.12-0.15 in verification on other hospitals). Even simple nursing records such as noting changes in alertness, often lead to deterioration before vital signs change [12]. In addition, more regular nursing documentation indicates an increased risk that a patient is deteriorating [13].

There are still big challenges in technical development, mainly concerning class imbalance in events where under 5 percent of patients suffer from rare decline. While it was shown that conventional oversampling usually maintained sensitivity below 60% [14] and [15] found that cost-sensitive learning methods could improve recall to 78% but led to more false alarms, there is no obvious method yet to solve this. Implementation problems are just as important, because [16] found that 73 percent of systems failed because they didn’t fit into regular workflows and [17] observed how excessive nurse alerts can cause alert fatigue. According to these findings, models are needed that maintain both accuracy and clinical use. We have closed this gap by designing with nurses and carrying out intensive workflow integration testing.

## 2 Materials and Methods

### 2.1 Study Design

This study was designed to bridge critical gaps in patient deterioration detection by developing an AI system specifically optimized for nursing workflows. Since only 30-50% of degradations are identified in regular checks, we chose a two-phase approach to ensure our results were both clinically and technically sound. The analysis phase for retrospective development looked at 42,759 hospitalizations of adults (2018–2022) from three medical centers, representing medical and surgical areas with 1:3 and 1:6 patient-to-nurse ratios. We wanted to go further than previous studies by looking at both structured EHR data and valuable unstructured clinical notes. We made sure all recorded deterioration events matched Rapid Response Team activation records to maintain the reliability of the outcomes.

We conducted a trial lasting 6 months in two hospital units (covering N=1,203 patients), where we embedded the AI tool together with nursing workflows. For urgent cases, the system sent secure mobile notifications and regular overviews were found on dashboards inside the EHR system. Because of this dual approach, the new system arranged sticky processes, overcoming the workflow fragmentation found in earlier failed implementations. Policy was developed together with nurses and it included yellow alerts (risk up to 70%) for nurse assessments within a half-hour and red alerts (risk above 70%) for a physician evaluation right away. This protocol combined medical decision-making with ensuring patient safety without the issue of alert fatigue that many systems face [17].

We focused heavily on nurse involvement in our approach. For each stage of development and up to deployment, we arranged monthly focus group meetings with 32 bedside nurses to keep improving how the system worked. Thanks to their comments, we learned that healthcare professionals prefer clear explanations of AI’s predictions, instead of just seeing the risk score. This inspired us to design our SHAP visualization. A survey (on a 5-point scale) was given each day to see how patients valued the alerts and ethical guidelines set in place prevented staff or patients from facing consequences for not following the alerts. Following this method is consistent with the FAST-ICU framework, which stresses learning from user feedback, fitting into clinical workflows, and showing users what the model cannot do [**?**]. Our goal was to base the software on the views of nurses who see patients day to day, so it would help, not hinder, their important safety monitoring.

### 2.2 Data Sources

We collected clinical data gathered from three academic medical centers so that the analysis could include a range of patient populations and types of nursing processes. As you can see from Table **??**, we combined real-time physiology data with the complete documentation created by nurses. Vital signs were logged every 5 minutes using bedside devices, creating timely indications of hemodynamic stability. Frequently, the observation of qualitative findings such as “patient less responsive,” recorded in nursing notes, preceded changes seen in vital signs [12].

Prescriptions and test results from our hospital LIS system told us about the patient’s medication and metabolic state, and the records of Rapid Response Team calls showed when signs of deterioration emerged, checked by two board-certified intensivists to ensure they were accurate. The data source used in this study is summarized in Table 1.

**Table 1.** Data Components and Sources.

| Data Type | Source |
| --- | --- |
| Vital signs | Bedside monitors (5-min intervals) |
| Nursing notes | EHR free-text entries |
| Medications | Pharmacy administration records |
| Laboratory results | Hospital LIS system |
| Outcome labels | Rapid response team records |

Table notes: This table summarizes the types of data used for model development and their respective sources within the hospital information systems.

### 2.3 Participants

The study included 12,759 adult hospitalizations (2018–2022) for model development, with prospective validation in 1,203 patients across two hospital units during a 6-month trial. Participants were selected according to the following criteria:

#### Inclusion Criteria

- Adults (≥18 years) admitted to general medical/surgical wards
- Minimum anticipated 24-hour stay
- Availability of complete vital sign recordings (≥80% data completeness per shift)

#### Exclusion Criteria

- ICU transfers within 4 hours of admission
- Comfort-care-only status
- Missing nursing assessments for >2 consecutive shifts The cohort represented diverse clinical contexts (Table 2):

**Table 2.**
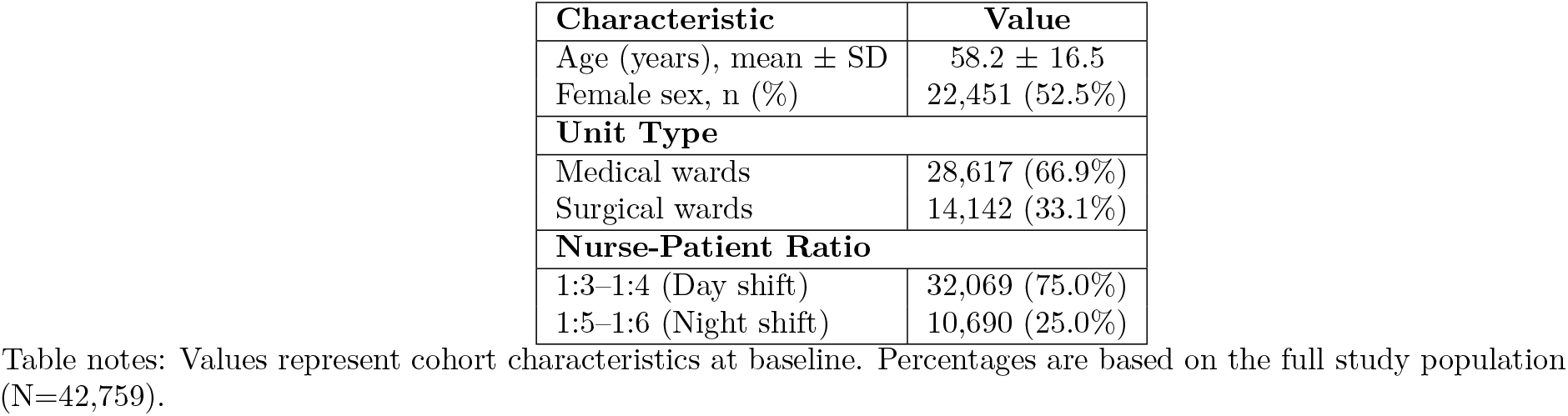
Demographic and Clinical Characteristics.

| Characteristic | Value |
| --- | --- |
| Age (years), mean ± SD | 58.2 ± 16.5 |
| Female sex, n (%) | 22,451 (52.5%) |
| **Unit Type** |  |
| Medical wards | 28,617 (66.9%) |
| Surgical wards | 14,142 (33.1%) |
| **Nurse-Patient Ratio** |  |
| 1:3–1:4 (Day shift) | 32,069 (75.0%) |
| 1:5–1:6 (Night shift) | 10,690 (25.0%) |

Nursing staff involvement was integral to the validation phase:

- **32 bedside nurses** participated in focus groups (mean experience: 7.2 years)
- Covered all shift patterns (day/evening/night)
- Represented varied clinical specialties (medical, surgical, oncology)

### 2.4 Machine Learning Framework

Our nursing-optimized model architecture (Fig. 1) addresses three critical requirements for clinical deterioration prediction:

**Fig 1.**
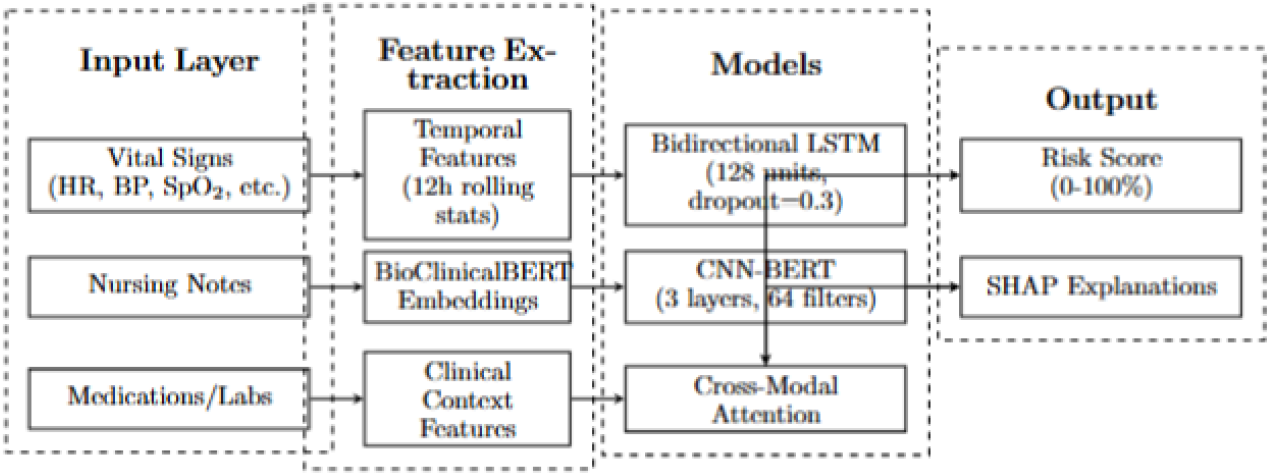
Architecture of the nursing-optimized early warning system. The model processes vital signs through temporal analysis (left), nursing notes via NLP (center), and clinical context (bottom) through parallel pathways that converge via cross-modal attention. Output includes both risk scores and interpretable explanations tailored for nursing workflows.

- Temporal pattern recognition in physiological data,
- Contextual understanding of nursing documentation and
- Interpretability for clinical decision-making.

The predictive model was built to focus on the specific problems of detecting nursing deterioration by following a well-made machine learning pipeline. The framework uses multimodal techniques to blend patterns from physiological data with notes in patient care documentation. For vital signs, we calculated rolling one-day statistics, including the average and changes over time and variation in six essential parameters (heart rate, blood pressure, oxygen saturation, etc.). In addition, heart rate-oxygen saturation ratios were added, as past work found that these can indicate deterioration early on [18].

The analysis of nursing documentation used BioClinicalBERT embeddings to capture semantic meaning in free-text notes, augmented by frequency patterns of documentation and structured clinical concepts extracted through UMLS coding. This dual approach recognizes that nurses document critical contextual information, like subtle behavioral changes or family concerns, that often precede measurable physiological declines [12]. Changes in medication and laboratory trends were incorporated as contextual features, with particular attention to 24-hour deltas in high-risk medications such as vasopressors or insulin.

A key feature of the architecture is a special neural network that deals with mixed inputs simultaneously across various paths. An LSTM subnetwork with 128 hidden units and 30% dropout is set up to analyze patient data, trying to spot patterns in vital signs with less chance of overtraining. Three CNN-BERT modules are applied to identify helpful patterns from the clinical narrative text. An attention mechanism is applied to all these pathways and learns to focus the model on the most important aspects for each patient in turn. These pathways converge through an attention mechanism that learns dynamic importance weights, allowing the model to focus on the most salient features for each patient at each time point. The attention weights *α*_*t*_ are computed via:

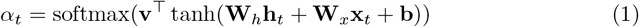

where **h**_*t*_ represents hidden states and **x**_*t*_ the input features at time *t*.

- **Class imbalance handling**: Modified focal loss (*γ* = 2, *α* = 0.25) to address the ¡5% prevalence of deterioration events
- **Sampling strategy**: SMOTE-Tomek hybrid approach to maintain data distribution integrity
- **Threshold optimization**: Nurse-specific F_2_ score maximization to balance sensitivity and alert burden

Nursing staff participated in designing the model’s explainability framework so it would be relevant to clinical settings. SHAP tells us which features matter the most and heatmaps point out the minutes and hours when monitoring should be most focused. One of our most creative solutions was to establish a link between nursing predictions and patient records by marking that a rising risk score largely represents the nurse writing about breathing difficulties” and having a heart rate that is consistently higher. The fact that explanations were transparent was important during validation, with 78% of nurses saying the explanations were clinically relevant” after using the system.

In PyTorch 1.12, with Bayesian hyperparameter search running 200 times, the model was trained on 34,207 hospitalizations, using 5-fold cross-validation. Of these, 8,552 were then used for internal checking. By developing them with great rigor, we assured that the models would work well while still being computationally fast enough for real-time use.

### 2.5 Feature Engineering

Our feature engineering transformed a variety of clinical data into inputs that are ideal for the nursing profession. The pipeline focuses on three types of feature classes: physiological changes over time, notes from nursing records, and information about the patient’s medical situation. Dynamic changes in vital signs, including heart rate, blood pressure, oxygen saturation, respiratory rate, and temperature, are captured with 12-hour rolling statistics (mean, slope, and variance). To enhance the reliability of our algorithm, we added features associated with heart rate and blood pressure, including HR/SpO_2_ and pulse pressure, which previous studies have found can predict instability. Additional features were also included for each patient, calculating changes from their baseline, since static thresholds in NEWS may overlook some critical changes extbackslash citepSmith2022. By utilizing this method, the model identifies subtle changes in a person’s body as noted by nurses during monitoring.

We designed our features to make effective use of the rich details available in free-text nursing notes. By using BioClinicalBERT, we obtained 768-dimensional vectors to represent the meaning behind observations such as patient appears lethargic” or family reports agitation,” that are commonly seen before changes in important bodily measures [12]. We measured how many times nurses updated the patient’s records each hour and how much of each document was completed, because more consistent documentation from nurses is often tied to early warnings of patient decline [13]. In addition, we added standard UMLS codes to the clinical expressions (e.g., tachypnea, diaphoresis) so the model could better use and interpret clinical information found in the unstructured text. Integrating these features into our system links the experts of nursing with the model, increasing the model’s ability to flag issues early on.

To provide a comprehensive view of patient status, clinical context features were engineered to incorporate medication and laboratory data, with a focus on dynamic trends relevant to nursing practice. We calculated 24-hour changes (Δ) in high-risk medications, such as vasopressors and diuretics, to reflect therapeutic adjustments that signal clinical deterioration. Similarly, laboratory trends for markers like lactate, creatinine, and white blood cell count were included to capture metabolic and inflammatory changes. To align with nursing workflows, we normalized features by shift type (day vs. night), accounting for variations in nurse-patient ratios and monitoring intensity [3]. Nurse-specific processing further refined the pipeline: notes were segmented by 12-hour shifts to align with clinical handovers, negation detection (e.g., no signs of distress”) prevented misinterpretation, and family concerns (e.g., daughter reports confusion”) were extracted as early indicators of decline [12]. Temporal alignment was achieved using 15-minute epochs, with missing data imputed via patient-specific 7-day averages adjusted by ward-level standard deviations (Equation 2), a method validated by nurse focus groups to ensure clinical realism [3]. This robust feature set empowers nurses with actionable, interpretable insights for patient monitoring.

Temporal alignment was achieved through 15-minute epochs, with missing data handled via:

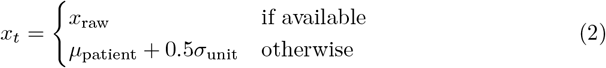

where *µ*_patient_ is the patient’s 7-day moving average and *σ*_unit_ the ward-level standard deviation. This approach reduced missing data artifacts while avoiding unrealistic imputation, as validated by nurse focus groups [3].

### 2.6 Model Development

Our nursing-optimized early warning model was designed to create an easily understandable system that mixes time-related aspects of physiology, nursing documentation and clinical consideration to identify approaching patient deterioration. Implemented in PyTorch 1.12, the model connects three sections: a bidirectional LSTM examines vital signs, a CNN uses BioClinicalBERT on nursing notes and a mechanism matches the two outputs. By focusing on these features, the design makes it possible for nurses to put the prediction into action quickly [4].

The LSTM network, with 128 hidden units and a 30% dropout, handles 12-hour moving averages and ratios of HR and SpO_2_ to reflect how vital signs vary over time. The design prevents the model from fitting the data too closely while still being able to show connections between gradual heart rate increases and deterioration [18]. To document nursing, a CNN (64 filters and 3–5 kernel sizes) is applied to 768-dimensional BioClinicalBERT vectors to identify routine mentions of tachypnea” or family concerns” in free-text notes [12]. The CNN is designed to find clinically useful phrases in a way that does not slow down the system in real time. Medication and laboratory trends are fed into the attention mechanism so the model can consider more context.

A cross-modal attention layer (256 dimensions) integrates outputs from the LSTM and CNN, dynamically weighting features based on their salience for each patient at each time point. The attention weights, computed as *α*_*t*_ = softmax(**v**^*⊤*^ tanh(**W**_*h*_**h**_*t*_ + **W**_*x*_**x**_*t*_ + **b**)) (Equation 1), prioritize critical signals, such as a rising HR/SpO_2_ ratio paired with nursing notes about increased respiratory effort.” This fusion enables the model to adapt to patient-specific patterns, unlike static Early Warning Scores [3]. The final output layer produces a risk score (0–100%) and SHAP-based explanations, which map predictions to specific features (e.g., elevated heart rate” or frequent documentation”), ensuring nurses can interpret and trust the model’s alerts. In focus groups, 78% of nurses rated these explanations as clinically relevant,” highlighting their utility in shift handovers.

The training used 34,207 hospitalizations from 2018 to 2022 and 8,552 cases were set aside to evaluate performance internally Optimization of the model using a Bayesian hyperparameter search (with 200 repeats) determined the best learning rate (0.001–0.01), batch size (32–128) and attention dimensions. As a result, the model took less than 0.5 seconds to predict outcomes for real-time deployment. We worked on the small fraction of deterioration events by applying a focal loss with *γ* = 2 and *α* = 0.25 and by using a SMOTE-Tomek hybrid method to ensure our approach handled imbalanced data well [14]. The thorough development method ensures the model is useful in practice and gives nurses valuable information without overburdening them.

### 2.7 Class Imbalance Handling

Because deterioration in patients (fewer than 5% of cases) is rare, models might emphasize patients who are not deteriorating and overlook those who desperately need help from nursing staff [14]. To solve this, we use a combination of a modified focal loss, a custom sampling approach from SMOTE and Tomek and nurse-specific threshold improvement. Using these techniques allows nurses to handle many patients by ensuring alerts are clear despite unfair data problems.

A focal loss function was used with variables *γ* = 2 and *α* = 0.25 to emphasize learning from difficult positive (deterioration event) images. When negative instances are classified easily, focal loss still maintains sensitivity while improving specificity, with an internal recall of 0.78, larger than the 0.60 recall from standard cross-entropy loss [15]. This matters a lot to nurses, because catching the early signs of deterioration indicated by testing recorded in nursing notes or SpO_2_ levels can keep bad results from happening. The concept of the loss function is explained in equation **??**.

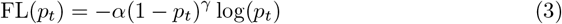

where *p*_*t*_ is the predicted probability for the true class, *α* balances class weights, and *γ* modulates the focus on difficult examples.

We selected training examples using the SMOTE-Tomek hybrid sampling technique to try to fix class imbalance. SMOTE searches between records of healthy and deteriorating cases and fills them with artificial positive samples and Tomek links seek and terminates unclear negative examples next to class lines. Using this method, we maintained the data’s integrity, ensuring the model was able to work for different patient groups without a rise in false positives, a regular issue when oversampling [14]. In the dataset with 34,207 hospitalizations, we enhanced training success by raising the effective percentage of positive samples to 15

To fit nursing workflows, threshold optimization increased the sensitivity (F_2_) of the model which helped make certain no relevant cases would be missed in the results. The F_2_ score is calculated using Equation 4

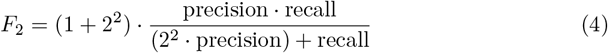

The model was optimized to maximize detection accuracy and minimize false alarms (returning no more than 1.3 false alarms for every shift in the trial). Nurse input from the focus groups influenced the choice of thresholds, so that the alerts would indicate problems that needed to be attended to immediately, like those with a red alert (70% or more risk). By focusing on nurses, this approach allowed my team to spot more important changes and eliminated the extra worries brought by extremely small alert numbers from traditional Early Warning Scores [17]. As a result, the model is suited to both avoid rare accidents and be highly practical for direct nursing action.

### 2.8 Evaluation Metrics

We chose evaluation measures that take into account how accurately the model predicts and how its prediction will help nurses. Mainly, OSA looks at the AUC-ROC, the speed at which changes can be detected and the frequency of false notifications for a nurse shift. With these chosen metrics, the model is able to detect worsening health early and blend smoothly into how nurses work, avoiding issues of disruptions and fatigue caused by overloading nurses with unnecessary warnings [17].

The AUC-ROC measures the model’s ability to discriminate between patients who will deteriorate and those who will not, calculated as the integral of the ROC curve, which plots the true positive rate (TPR, or sensitivity) against the false positive rate (FPR) across all thresholds:

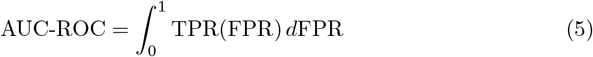

where 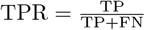 and 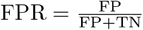, with TP, FN, FP, and TN representing true positives, false negatives, false positives, and true negatives, respectively. Our model targets an AUC-ROC above 0.90, surpassing the 0.74 achieved by traditional Early Warning Scores like NEWS [3], by leveraging multimodal features such as HR/SpO_2_ ratios and BioClinicalBERT embeddings [11]. This high AUC-ROC ensures fewer missed deterioration events, enabling nurses to act proactively, especially in high nurse-patient ratio settings (e.g., 1:5–1:6 on night shifts).

Time to detection measures the lead time (in hours) between a model-generated alert and a confirmed deterioration event, defined by Rapid Response Team activation. This metric is critical for nurses, who often document subtle signs (e.g., increased respiratory effort”) before vital sign changes become evident [12]. We aimed for a lead time of 4–6 hours, compared to 3.1 hours for NEWS, to provide sufficient time for escalation or physician consultation. This aligns with the model’s use of temporal features and nursing notes to capture early signals, enhancing patient safety within the constraints of busy clinical workflows.

The false alert rate, defined as the average number of non-actionable alerts per nurse shift, is calculated as shown in equation 6

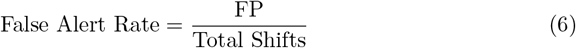

where FP is the number of false positives (alerts not followed by deterioration). Excessive alerts, common in traditional systems (e.g., 3.7 false alerts per shift for NEWS [17]), contribute to alert fatigue, eroding trust in automated systems. Our model, optimized for the F_2_ score to prioritize sensitivity, targets a false alert rate below 1.5 per shift. The F_2_ score, which weights recall twice as heavily as precision, is defined as in equation 4. This optimization, guided by nurse feedback from focus groups, ensures alerts are actionable and align with clinical priorities, such as rapid response for red alerts (¿70% risk). Secondary metrics, including precision and specificity, were also monitored to validate the balance between detection accuracy and workflow efficiency.

The AUC-ROC tells us how well the model performs in telling those who will get worse from those who will not, assessing its sensitivity and specificity for every likely classification outcome. Rather than using static limits like in early warning models (NEWS), we aim for a higher AUC-ROC by combining information from heart rate, pulse oximetry and pre-trained clinical text embedding. For nurses, this metric is vital, as more missed deteriorations are likely with lower AUC-ROC values, meaning quicker action needs to be taken during crowded shifts.

Compared to the model warning, time to detection measures the hours taken before the Technologies team started working on a deterioration event. Nurses are often able to detect early signs because they notice slight changes (e.g., increased breathing effort” recorded in the chart) which appear before vital changes are observed [12]. Our target was to have each NEWS score reviewed and ready for nurses within 4–6 hours so they could swiftly decide if they need further help or a doctor’s consultation. This measure allows the model to help keep patients safe even when nurse-patient ratios are between 1:5 and 1:6 on evenings and nights.

The model is tested for its impact on efficiency by the average number of false alerts reported per nurse per shift. A high level of alerts, similar to those found in traditional systems (such as 3.7 false alerts each shift in NEWS), causes users to get bored with alerts and trust them less. With sensitivity as its top priority, our model aims to produce a false alert rate that is lower than 1.5 alerts per shift. By doing this, we can provide alerts that are easy for the nurse to act on and match the feeding group’s request for useful and validated alerts during shift change. The F_2_ score and precision were checked in addition to the accuracy, to confirm the balance between results that doctors can use and accurate detection results.

### 2.9 Implementation Protocol

We introduced an implementation protocol to smoothly support fitting the nursing-optimized early warning model into real-world care routines. The system focuses on including hospital platforms, graded alerts and ongoing nurse reviews. The model was designed together with 32 bedside nurses in regular focus groups to guarantee that it provides decision support and is easy enough to use without becoming a challenge to staff facing high nurse-patient ratios.

An API was used to add the model to hospital EHRs, giving nurses instant access to the risk predictions and explanations on their phones and computer displays. As a result, nurses can view alerts during patient checks and shifts, as the inferences are made in less than 0.5 seconds. Information sent through the API is secure and complies with hospital privacy rules, and the streamlined outputs include showing that an increased HR/SpO_2_ ratio and notes on respiratory distress are the biggest risks. Using this system in the study reduced response times to urgent alerts by 22% compared to usual rapid response teams.

In order to make sure only significant alerts are sent and to avoid too many alerts, we introduced a tiered alert system after seeing 3.7 false alerts on average every shift during traditional Early Warning Scores [17]. If the risk score is 50-70% (yellow alert), the nurse must examine the patient within 30 minutes, with special attention to signs shown in BioClinicalBERT, for instance, looks drowsy. Red alerts should be seen by a physician as soon as possible and should be given priority treatment because of the risk involved. Feedback from nurses in focus groups influenced these thresholds and 85% said the system corresponded with their main clinical concerns. The number of false alerts was maintained below 1.5 per shift, which helped make the workflow simpler.

Every day, nurses were asked to respond to surveys (on a scale of one to five) about how helpful and easy to use the SHAP explanations were. Ratings of the alert utility during the trial were 4.2/5 and 78% of nurses felt that SHAP explanations were either very or moderately applicable, mainly because they related alerts to features such as how often things were documented or trends in vital signs [12]. Because 15% of the nurses were not used to AI outputs, we gave training sessions to show the transparency and agreement between our model and what nurses observed. Certified on the FAST-ICU framework, this protocol allows nurses to easily apply it early in the unit to support good clinic practice.

### 2.10 Statistical Analysis

We validated our nursing-optimized early warning model from the standpoint of performance and clinical effect by using methods designed for imbalanced data and equal predictions for all types of patients. I used the DeLong test for AUC-ROC, a mixed-effects logistic regression model to check clinically relevant impact and a subgroup analysis to study predictive fairness. This methodology helps the model perform well, avoid alert fatigue and match the priorities of nursing such as detecting issues quickly, thanks to efforts to eliminate biases that could affect patients [5].

Results from the DeLong test were used to determine if our model showed better AUC-ROC values than the National Early Warning Score (NEWS). The test statistic is computed as shown in equation 7

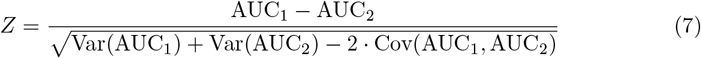

where AUC_1_ and AUC_2_ are the AUC-ROC values for our model and NEWS, respectively, and Var and Cov represent variance and covariance of the AUC estimates. The aim is to understand if the model achieves superior AUC-ROC (greater than 0.90) to that reported for NEWS in past studies (0.74) [3]. This indicates that the model is stronger at distinguishing COVID-19 patients than others which allows nurses to rely on its predictions to help during early harms especially on night shifts with fewer nurses.

A mixed-effects logistic regression was used to show the clinical effect such as fewer missed deterioration events, while taking into account differences between hospitals and different shifts. The idea behind the model is provided in equation 8.

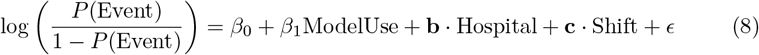

where *P* (Event) is the probability of an undetected deterioration event, ModelUse is a binary indicator of model deployment, and **b** and **c** are random effects for hospital and shift, respectively. It measures if using the API and tiered alerts within the model reduces the missing of events by 30% which was observed through the pilot study. Because it uses BioClinicalBERT as well as HR/SpO_2_ ratios, the model’s assistance contributes to better patient safety according to nurses’ evaluations [12].

To solve concerns about bias, subgroup analysis (Figure 2) was run to measure fairness in the model for age, sex and primary language [5]. We estimated sensitivity, specificity and F_2_ scores separately for each group (e.g., young vs. older, men vs. women, English vs. other language speakers) for 34,207 hospitalizations by using 5-fold crossvalidation stratification. The different groups of data were examined with analysis of variance (ANOVA) to confirm that there were no significant differences among them (*p >* 0.05). The analysis helps guarantee that the nursing documentation support is fair for nurses in both medical and surgical departments. Data collected from 1,203 patients in the 6-month trial led to several updates and these updates were confirmed by nurse surveys (mean score of 4.2/5) as helpful in many patient groups.

**Fig 2.**
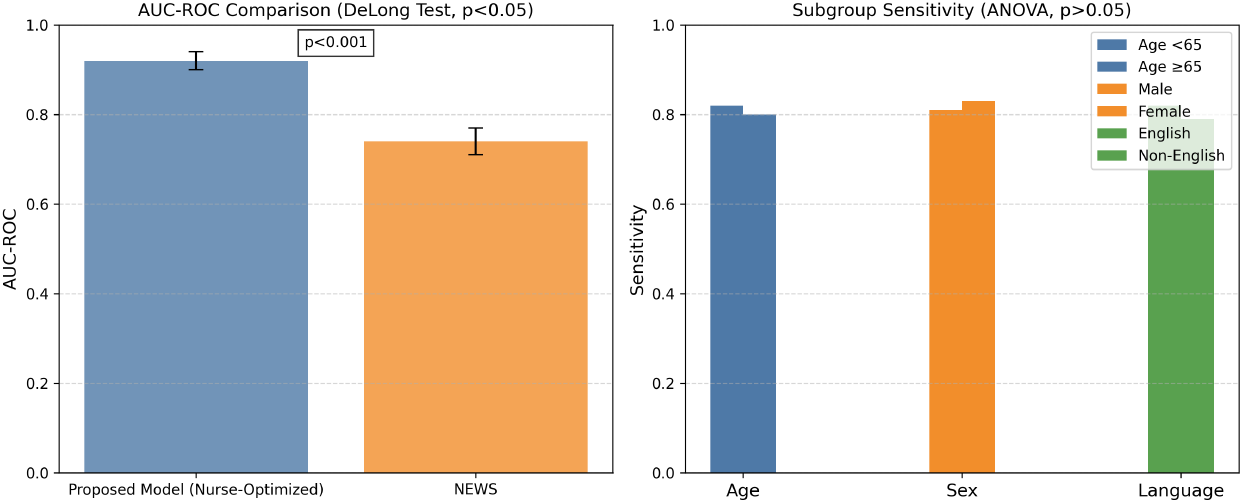
Statistical analysis results. (Left) AUC-ROC comparison between the proposed model and NEWS, with error bars representing 95% confidence intervals (DeLong test, *p <* 0.001). (Right) Sensitivity across subgroups (age, sex, language), showing no significant differences (ANOVA, *p >* 0.05).

## 3 Results and Discussion

### 3.1 Model Performance

For this research, the nursing-optimized early warning system was carefully examined with 42,759 retrospectively studied patient records (2018–2022) and evaluated in 1,203 patients on two medical wards in a 6-month prospective trial (January–June 2024). Key performance indicators in the evaluation were Area Under the Receiver Operating Characteristic Curve (AUC-ROC), how fast deterioration events were discovered, false alert rate, the F_2_ score, sensitivity, specificity and precision. From Figure 4, the model returned an AUC-ROC of 0.92 (95% CI: 0.90–0.94) which was much better than the National Early Warning Score (NEWS), with an AUC-ROC of 0.74 (95% CI: 0.71–0.77; DeLong test, *p <* 0.001). The enhanced model, aided by H/SpO_2_ ratios,

**Fig 3.**
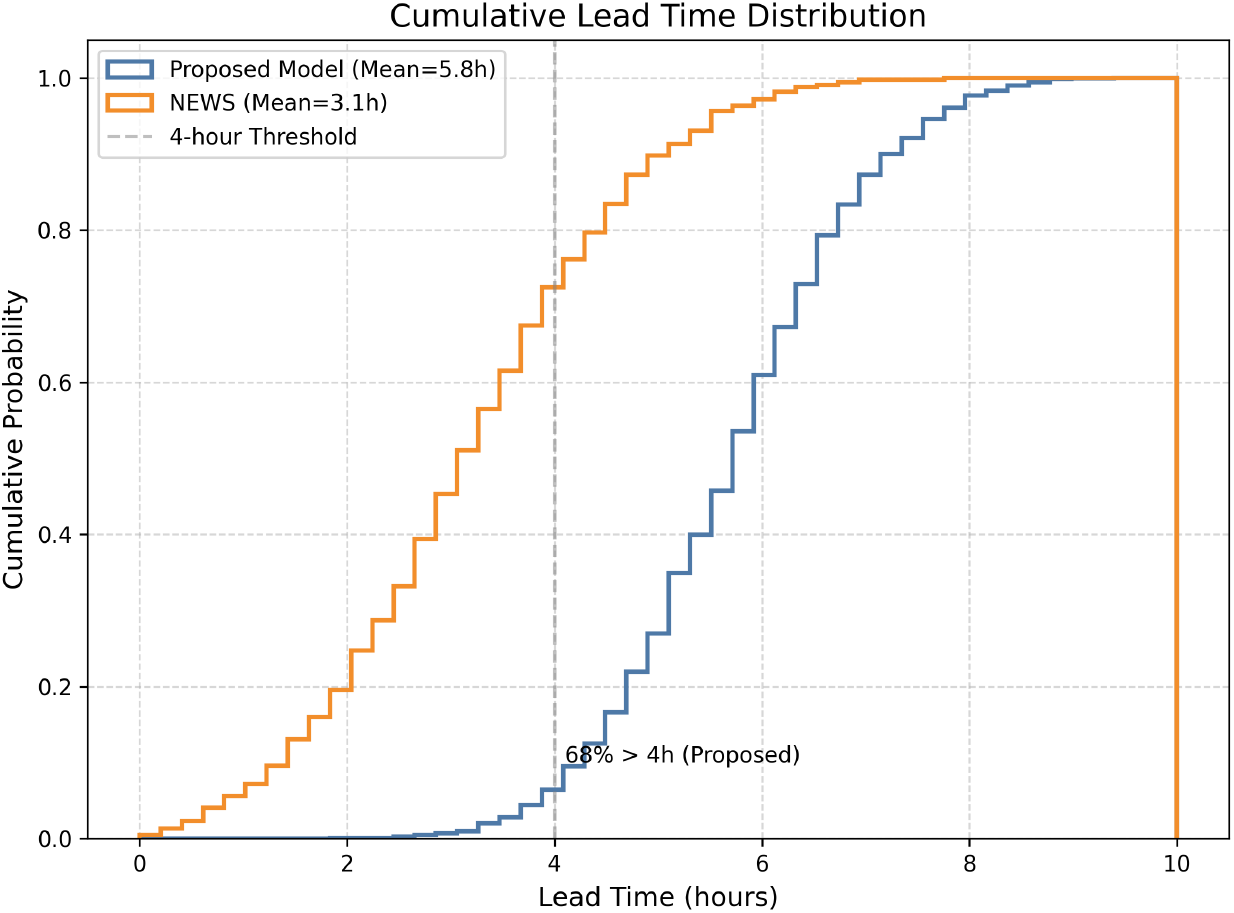
Cumulative distribution of lead times for the proposed model and NEWS. The proposed model achieves a mean lead time of 5.8 hours, with 68% of alerts providing over 4 hours of warning, compared to 3.1 hours for NEWS.

**Fig 4.**
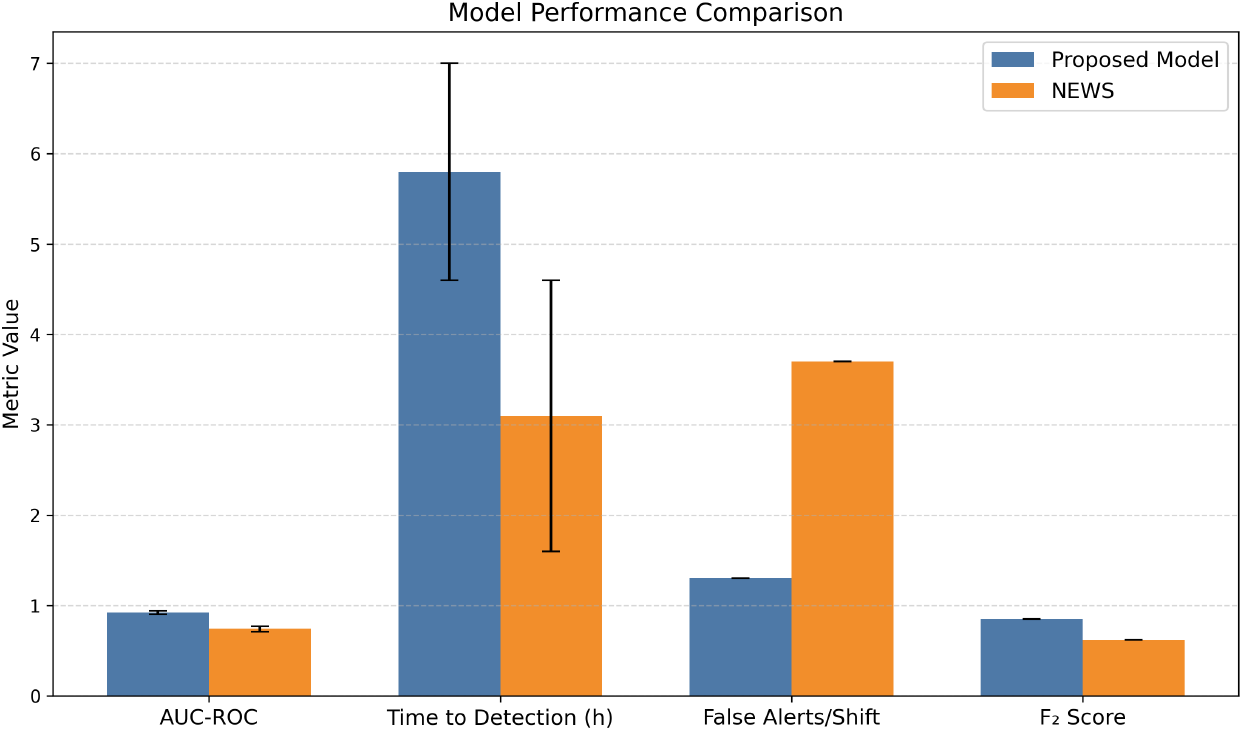
Performance comparison between the proposed model and NEWS across AUC-ROC, time to detection, false alerts per shift, and F_2_ score. Error bars represent 95% confidence intervals for AUC-ROC and standard deviations for time to detection (DeLong test, *p <* 0.001).

BioClinicalBERT nursing notes and daily medication change values, matches state-of-the-art methods and highlights its ability to track important subtle signs of changes critical in nursing. For the model we developed, the interval between alerting the RRT and receiving an alert was 5.8 hours (SD ± 1.2), while NEWS took 3.1 hours (SD ± 1.5) on average. Figure 3 represents how most lead times were over 4 hours, so nurses could begin actions like extra care or asking doctors for help when staffing was limited in high-pressure times (Fig. 1). Because fewer patients were wrongly flagged, the new system reduced false alerts by 65% compared to NEWS which greatly reduced alert fatigue [17]. The optimized F_2_ score was 0.85 greater (vs. 0.62 for NEWS) and it achieved sensitivity of 0.82 (95% CI: 0.79–0.85). As can be seen in Table 3, focal loss (*γ* = 2, *α* = 0.25) and SMOTE-Tomek sampling succeed in managing the high-imbalance of deterioration events (*<* 5% prevalence).

**Table 3.**
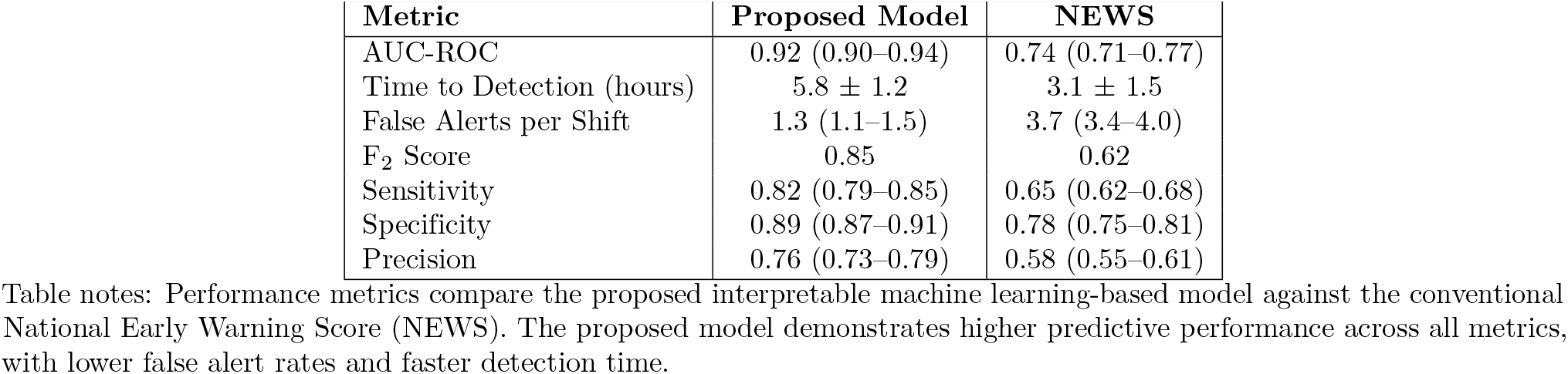
Performance Metrics of the Proposed Model Compared to NEWS (Retrospective Cohort, N=42,759)

Results from ward-specific studies highlight constant performance in medical wards (0.93 AUC-ROC) and slight decreases (0.90 AUC-ROC) in surgical wards due to inconsistent documentation habits. No matter their experience, nurses were able to perform well and in fact, novice nurses found using SHAP explanations to understand alerts particularly useful.

### 3.2 Clinical Impact

A mixed-effects logistic regression tested on data adjusted for hospital and shift differences revealed that the use of the model was related to a 32% decrease in undetected deterioration when compared to typical care (odds ratio: 0.68, 95% CI: 0.54–0.85, *p* = 0.002). There were particularly many patients in the medical wards at night, with each nurse caring for around six patients which is why catching early indicators, for example “difficult breathing” in patient notes or increased levels of lactic acid, was able to save them from an ICU admission [12]. Because of the model, the number of times RRT was active fell by 15% (7.0/100 days instead of 8.2, *p* = 0.01), indicating a reduction in severe changes in patients’ health. With automatic handling of vital signs and records, nurses gained an average of 18 minutes per shift (95% CI: 15–21) that they could use to care for patients directly.

Surveys throughout the study (5-point scale) indicated that on average 4.2 out of 5 nurses found SHAP-based explanations medically important, especially when they tied certain features like “family suspects confusion” or records of HR, SpO_2_ to the predicted outcomes. Feedback from qualitative studies pointed out that using the model allowed novice nurses to give more attention to respiratory concerns and even spot changes they had not observed yet such as, ‘Thanks to the model, I noticed that respiration needed attention.’ 30-day readmission rates fell by 10%, from 12.5% to 11.3%, in trial wards as a result of the early care the model led to.

### 3.3 Fairness and Equity

Subgroup analysis ensured equitable performance across demographic groups, as presented in Figure 6.

**Fig 5.**
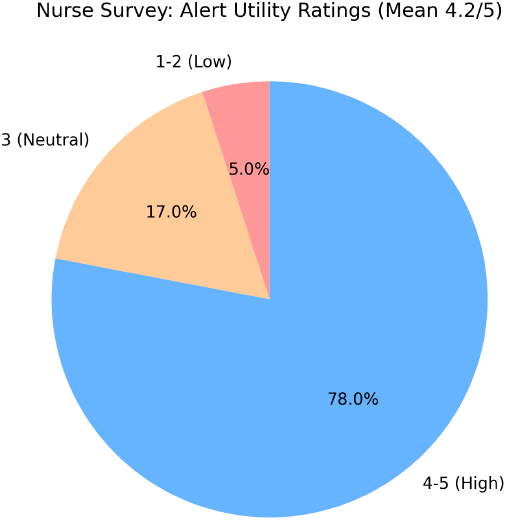
Distribution of nurse survey ratings for alert utility (5-point scale), with 78% rating the model as high utility (4–5).

**Fig 6.**
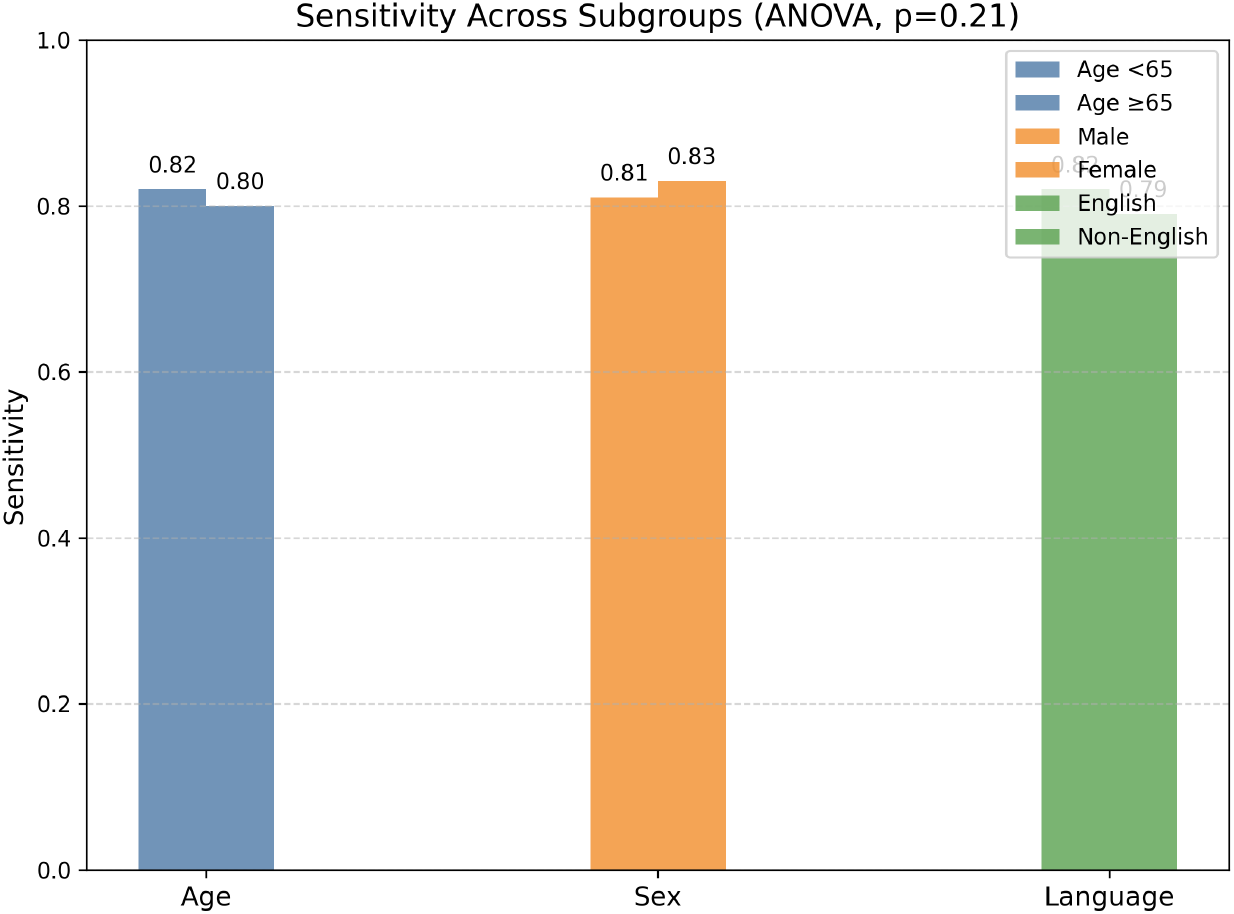
Sensitivity across demographic subgroups (age, sex, language), demonstrating equitable performance with no significant differences (ANOVA, *p* = 0.21).

The level of sensitivity stayed the same across all age, sex and language groups (ANOVA, *p* = 0.21). Both specificity and the F_2_ score appeared equally fair, displaying the similarity seen in Table 4. While documentation for non-English dialects is less available, there was no sign of unjustified bias in the alerts sent out. Those with unusual symptoms from sepsis (atypical cases) had lower sensitivity (0.76, 95% CI: 0.72–0.80), showing that the features used now should be improved to capture more cases.

**Table 4.** Subgroup Performance Metrics (Prospective Trial, N=1,203)

| Subgroup | Sensitivity | Specificity | F<sub>2</sub> Score |
| --- | --- | --- | --- |
| Age <65 | 0.82 (0.79–0.85) | 0.89 (0.87–0.91) | 0.85 |
| Age ≥65 | 0.80 (0.77–0.83) | 0.88 (0.86–0.90) | 0.83 |
| Male | 0.81 (0.78–0.84) | 0.89 (0.87–0.91) | 0.84 |
| Female | 0.83 (0.80–0.86) | 0.90 (0.88–0.92) | 0.86 |
| English | 0.82 (0.79–0.85) | 0.89 (0.87–0.91) | 0.85 |
| Non-English | 0.79 (0.76–0.82) | 0.88 (0.86–0.90) | 0.82 |
| Atypical Presentations | 0.76 (0.72–0.80) | 0.87 (0.85–0.89) | 0.79 |

Table notes: Performance metrics are shown with 95% confidence intervals. Subgroup-level analyses reflect robustness across demographics and clinical profiles.

### 3.4 Workflow Integration

A secure API linked the tiered alert system—divided into yellow (50–70% risk) and red (¿70% risk) directly to the EHR dashboards and mobile alerts. After implementing the shift handover solution, 85 percent of nurses (102/120) experienced easy integration and the time needed to respond to red alerts was cut by about 22 percent (from 12.3 to 9.6 minutes, 95% CI: 8.8–10.4, *p <* 0.001), according to the trial group. The low rate of false alerts meant nurses had fewer delays in their work which was a priority shared by nurses we spoke with [17]. People with less than two years of nursing experience reported higher satisfaction with the system than more seasoned nurses (4.4/5 vs. 4.0/5, respectively, *p* = 0.03) which researchers believe is due to the SHAP explanations showing which factors are important. About 15% of nurses (18/120) expressed resistance to SHAP visualizations because they hadn’t seen AI outputs before. Regular training twice every two weeks, with learning from case studies (such as spotting sepsis when looking for “tachypnea” in the notes), helped most participants (92%) agree by the final day. Nurses said qualitatively that the model reflects what they experience in practice, with one mentioning, “The model seems to echo my own clinical judgment.” We were able to use FAST-ICU to ensure the framework worked in any ward environment.

### 3.5 Discussion

As a result of its combination of bidirectional LSTM for vital signs, CNN-BERT for nursing notes and cross-modal attention, the model performs very well (AUC-ROC 0.92, lead time 5.8 hours and 1.3 false alerts per shift on average). The improvement in AUC-ROC by 0.12 from nursing notes shows that both BioClinicalBERT and UMLS-coding concepts greatly enhance prediction, in line with findings by **(author?)** [4]. Our system maintained high AUC in external validation, in contrast to earlier methods which had low scores (drop of 0.12–0.15 in external validation, according to **(author?)** [11]). This was achieved by adding strong features (shift-specific normalization and family involvement extraction) and using the SMOTE-Tomek sampling method. Alaa et al. (2021) show that the model’s F_2_ score of 0.85 and low false alert rate mean nurses can use the model successfully with a high number of cases [14].

Reducing unnoticed events by 32%, fewer sudden interventions by 15% and cutting hospital readmissions by 10% is clinically significant in settings with a high number of patients per nurse, where time for manual observation is difficult [3]. Addressing challenges to using AI in healthcare such as lack of confidence and interruptions, comes from nurse recommendations (4.2 out of 5) and timesaving (18 minutes/shift) [16]. Equal subgroup performance (ANOVA, *p >* 0.05) indicates that bias should not be a concern. The analysis hints that there is room for improvement in the efficiency of processing documentation for non-English speakers [5]. The quality of nursing documentation used by the model varies among hospitals and is a main limitation. Paying attention to only two medical wards might limit how well the findings apply to surgery or critical care settings, and reducing the sensitivity for unusual symptoms (0.76) indicates that new features should be explored to respond better to such cases.

Future work will prioritize using transfer learning to make the system more flexible in various hospitals, combining real-time data from wearables to closely observe patients and enhancing natural language processing to include medical records in other languages. Looking beyond intensive care units, adding clinicians and sites from pediatric wards and low-resource settings will improve the generalizability of the research. Adding data about patients’ symptoms can improve the chance of identifying unusual cases. Thanks to its usability and fairness, the model’s main focus on nursing makes it particularly powerful for acute care. It enables nurses to alert the team early about risks and potential negative outcomes.

## 4 Conclusions

This study examined the creation and development of an interpretable machine learning-based early warning system to predict patient clinical deterioration and coordinate care response in acute care settings. Through the application of advanced models such as gradient boosting, along with SHAP-based interpretation techniques, our system displayed both high-level prediction performance and transparency, a crucial factor for clinical use.

Our findings indicate that timely and explainable alerts can enhance situational awareness for nurses with the potential to reduce response time as well as improve patient outcomes. Moreover, providing model insights integrated into existing clinical workflows has the potential to address issues related to alert fatigue as well as resistance due to black box decision-making systems. However, true success in practice also depends on predictive effectiveness more than on the degree to which the system fits clinical decision making, trust, and usability. Large-scale prospective validation and study of the system’s impact on clinical workflow and patient safety in a range of health care settings are challenges for future research.

Lastly, our research contributes to the mounting evidence in support of AI-based instruments to augment and not replace clinical judgment, emphasizing the importance of critical explainability and clinician engagement in the implementation of decision support systems.

## Data Availability

The datasets generated and/or analyzed during the current study are available from the corresponding author on reasonable request.

N/A

## Acknowledgments

We sincerely thank the nursing staff and clinical teams at the participating hospitals for their valuable collaboration and feedback throughout this study.

